# Determinants of the plasma metabolome: cross-sectional and longitudinal associations over 6 years in the NESDA cohort

**DOI:** 10.64898/2026.01.14.26344096

**Authors:** Dennis Klose, Yuri Milaneschi, Laura K M Han, Nic J A van der Wee, Matthias Arnold, Christopher R Brydges, Siamak Mahmoudian Dehkordi, Gabi Kastenmüller, Rima Kaddurah Daouk, Brenda W J H Penninx

## Abstract

The plasma metabolome represents a valuable molecular readout of a person’s physiological state, yet its relation to health, stress and lifestyle remains underexplored collectively. Here, we conducted an untargeted metabolomics analysis using 3804 paired samples from 1902 participants of the Netherlands Study of Depression and Anxiety at baseline and 6-year follow-up, quantifying 680 plasma metabolites. We characterised five metabolome profiles using principal component analysis, three with distinct biochemical enrichments related to transmembrane transport, sphingolipid and amino acid metabolism. Metabolite levels showed moderate intrapersonal correlation between baseline and 6-year follow-up (ICC_median_ = 0.482), and only 22% of metabolites showed standardized mean differences greater than 0.2, indicating minimal 6-year population-level change. Multivariate linear modelling on 18 determinants across demographics, psychosocial environment, lifestyle, somatic and mental health explained a maximum of 35% of baseline metabolome profile variance and 12.2% in 6-year changes. Demographic (e.g., sex, age), somatic health (e.g., BMI, medication) and lifestyle factors (e.g., smoking, alcohol intake) demonstrated strong associations, while psychosocial factors and mental health contributed minor explained variance. Altogether, our study provides novel insights into the cross-sectional and longitudinal implications of health, stress and lifestyle exposures for the plasma metabolome, contributing to the understanding of metabolomic signatures in population studies.

## Introduction

The plasma metabolome represents the collection of exogenous chemicals and endogenous, small molecules originating from active metabolic processes, consisting of diverse compounds which facilitate energy uptake and storage, support cellular communication and maintain structural integrity (1). While several key influences on the blood metabolome composition have been identified in previous analyses using serum or plasma—including sex, age, genetics, diet, the gut microbiome, anthropometrics, circadian rhythm and diseases (2–6)—many sources of variation remain unexplored. Importantly, existing research has predominantly examined these influences in isolation, leaving a significant gap in our understanding of how multiple diverse factors contribute collectively. Metabolic alterations may play a fundamental role in the development of somatic and mental illnesses and can often be associated with environmental exposures such as medication, pollutants, lifestyle choices or psychological stressors (7–17). While the metabolome-wide influence of psychological stressors and mental health has been investigated in previous studies (3, 18), the role of more varied and detailed psychosocial factors has not been thoroughly investigated so far.

Here, we present the largest LC-MS-based study to date (n=1902) examining the associations of 18 determinants with 680 identified human plasma metabolites leveraging both cross-sectional and longitudinal data from 6-year follow-up assessments (FU6), overcoming limitations of previous studies pertaining to sample size, assessed determinants and timepoints. Our primary objective is to estimate the percentage of variance explained by determinants spanning five conceptual domains (demographics, psychosocial environment, lifestyle, somatic and mental health) in metabolite levels measured cross-sectionally or modelled as longitudinal change over 6 years. For this, we derive distinct metabolome profiles using principal component analysis and characterize them based on their potential enrichment in biochemical processes and subsequently use these to establish metabolome baseline scores and metabolome change scores for linear modelling. Altogether our study gives insight into which characteristics and life experiences are embedded into the plasma metabolome on two timescales. These findings may provide the basis for future research to develop personalised lifestyle interventions targeting psychosocial wellbeing or lifestyle-related behaviours to optimise metabolic health conditions.

## Results

### Metabolome principal components differ in their biochemical composition

To characterise the metabolome profiles of our study participants, we first performed a principal component analysis (PCA) on 3804 paired samples from 1902 subjects and 680 metabolite levels (**Figure 1**, **Suppl. Table 1**). We observed a shift of the FU6 samples compared to the baseline samples in the dimensionality-reduced metabolome space (**Figure 2a**). To enhance the relevance and interpretability of our findings, we retained PC1 to PC5 because these together captured the substantial majority (27%) of total variance between samples (**Suppl. Figure 1a**). Inspection of the scree plot demonstrated a clear elbow at PC5, after which additional PCs only contributed minimally (< 3% per PC) to the total explained variance. Beyond PC5, incremental increases were negligible and did not meaningfully improve representation of the underlying data structure. To validate the underlying PC structure, we conducted a separate PCA on 910 held-out baseline samples for which no follow-up measurement was available (**Suppl. Table 2**). We correlated the 680 metabolite loading scores from the first PCA (discovery) with those from the second PCA (validation) and determined moderate to high Pearson’s correlations for the respective loading scores (PC1: *r* = –0.94, PC2: *r* = 0.96, PC3: *r* = –0.94, PC4: *r* = –0.82, PC5: r = 0.58), confirming that the analysed dimensionality-reduced data captures meaningful and consistent information (**Suppl. Figure 1b**). Likewise, the explained variance per PC from the discovery PCA could be confirmed using the validation samples (**Suppl. Figure 1a**). When considering change over 6 years, PC1 and PC3 showed the strongest change, although of small size, between the two timepoints with standardised mean differences (SMD) of –0.28 and 0.25 (**Suppl. Figure 1c**). After conducting a PCA test, 604 out of 680 metabolites (88%) significantly contributed to PC1 scores, whereas 257 (37%), 232 (34%), 235 (34%) and 180 (26%) metabolites significantly contributed to PC2,3,4 and 5 scores, respectively (**Suppl. Table 3**). Unique or overlapping metabolites from all five PCs are shown in **Suppl. Figure 1d**. In addition to the broad 6-year changes observable in PCA, we assessed the stability of metabolite levels between the two timepoints individually: Population-level changes higher than absolute SMD > 0.2 were not common, shown by only 22% of metabolites (**Figure 2b**), with moderate within-subject correlation (ICC_median_ = 0.482). Interestingly, we observed a strong reduction in per– and polyfluoroalkyl substances (PFAS) over 6 years and a striking increase in argininate, dibutyl sulfosuccinate and sarcosine. Dehydroepiandrosterone sulfate and pregnenolone sulfate (neurosteroids known to decrease with age) were also among the metabolites with the highest reduction over 6 years. For detailed metabolite level differences between timepoints see **Suppl. Figure 2** and **Suppl. Table 4**.

**Figure legend 1.**
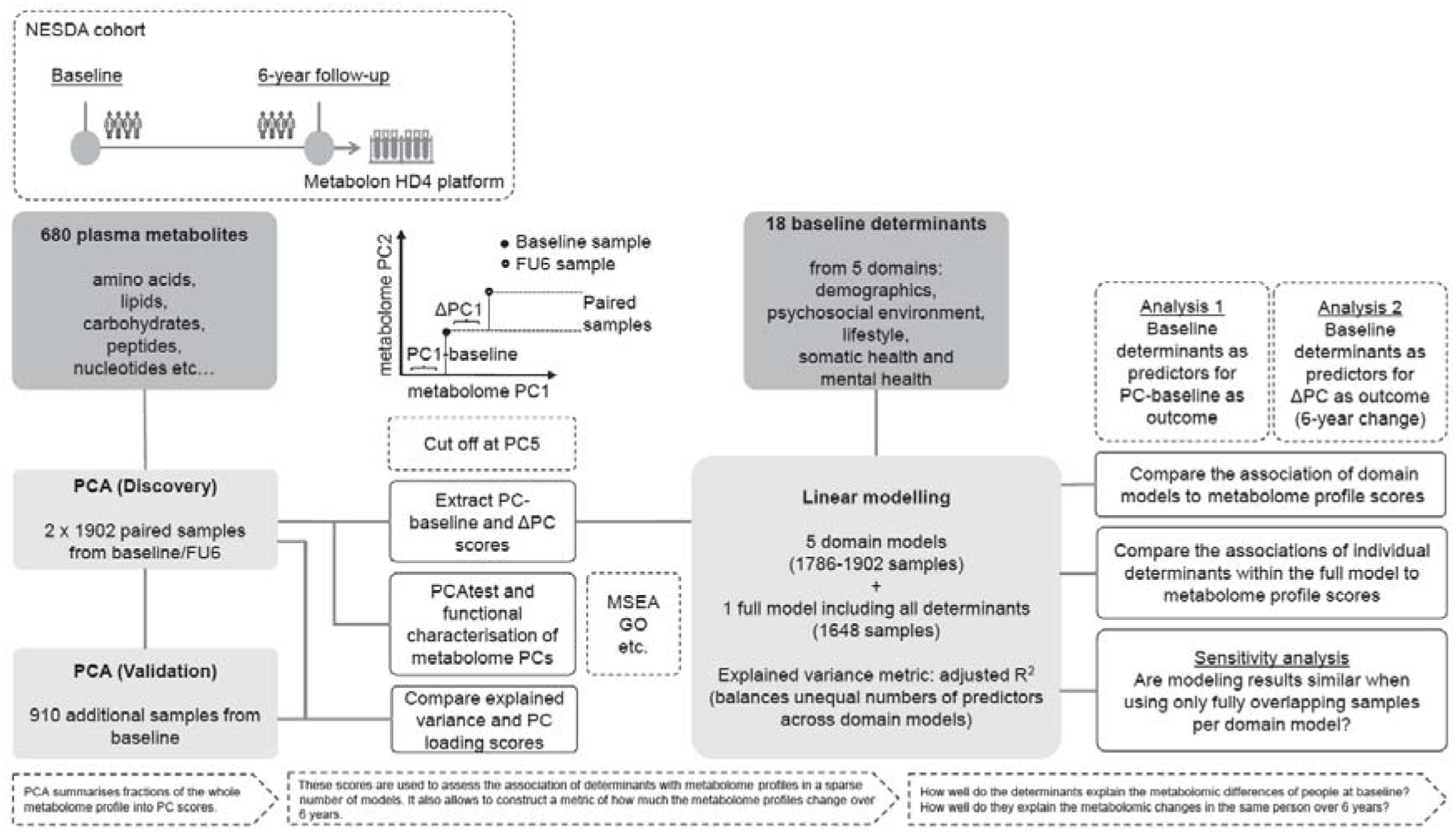
– Study design and workflow. PCA – Principal Component Analysis, FU6 – 6-year follow-up, MSEA – Metabolite Set Enrichment Analysis, GO – Gene ontology analysis.

**Figure legend 2.**
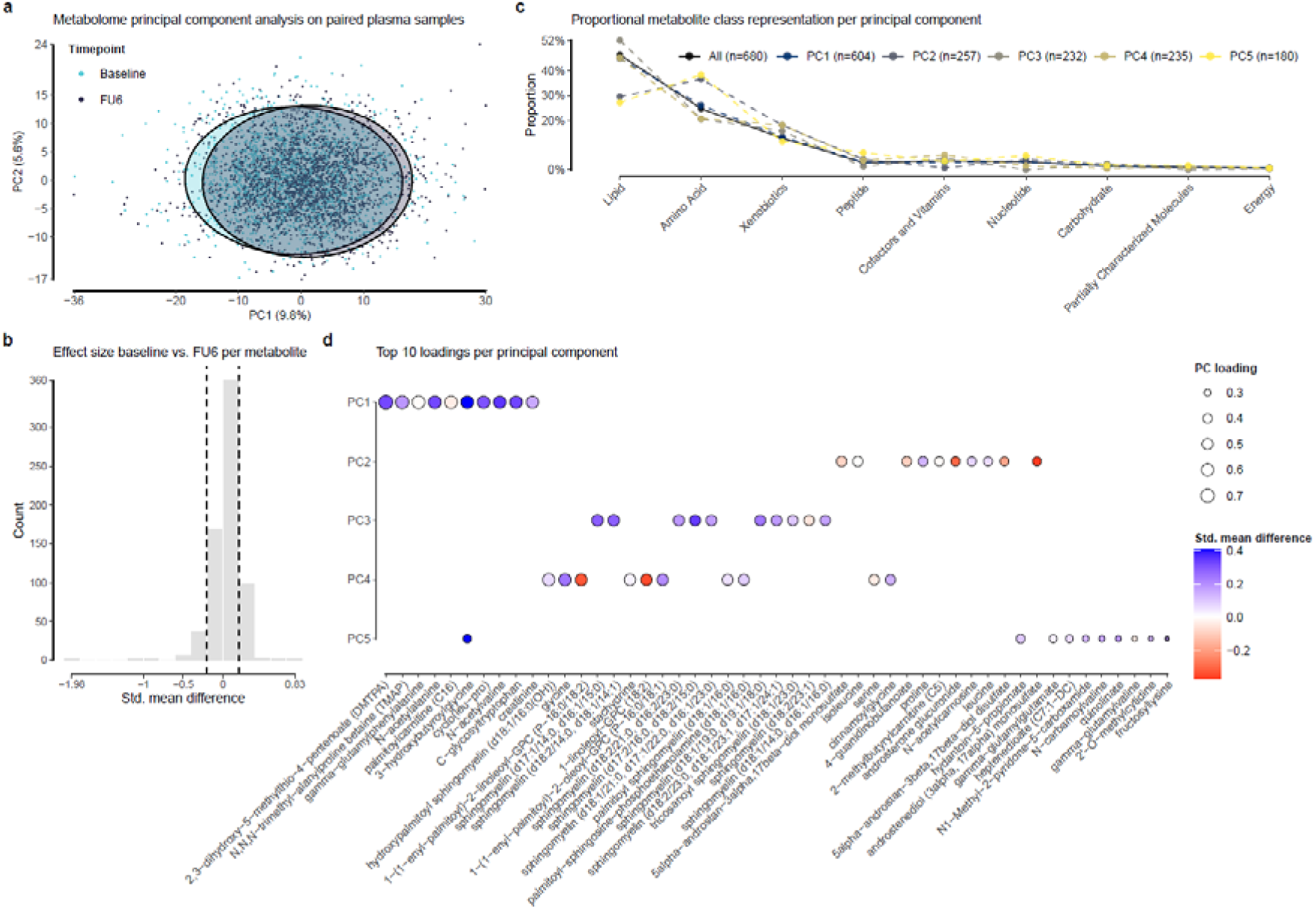
– Description of the first five metabolome PCs. **a** In a PCA plot using PC1 and PC2, the samples from baseline and 6-year follow-up (FU6) display a shifted metabolomic state (the two density circles do not fully overlap). **b** All 680 metabolites’ standardized mean difference (SMD) for baseline versus FU6 shown as a histogram. Dashed lines mark SMD = ±0.2. Count refers to number of metabolites. **c** After a PCA test, the significant loadings (metabolites) per PC are shown categorised by metabolite class. While PC1 metabolites do not show different metabolite class proportions compared to all measured metabolites, PC2-5 display divergent proportions for several metabolite classes, especially lipids and amino acids. **d** Metabolites with highest loading scores per principal component. A higher loading score indicates more importance for the respective PC score.

We next aimed to describe the significant metabolites per PC in terms of class and importance: While PC1 metabolites were a general reflection of the overall composition of measured metabolites, we further observed distinct metabolite proportions across the other components: PC2 was characterized by decreased lipids and cofactors/vitamins alongside increased amino acids and xenobiotics. PC3 showed an enrichment in lipids and xenobiotics but depletion in amino acids, peptides, and nucleotides. PC4 exhibited reduced amino acids yet elevated xenobiotics and cofactors/vitamins. Finally, PC5 displayed lower lipid content while showing higher levels of amino acids, peptides, and nucleotides. (**Figure 2c**). The top ten metabolites with the highest loading scores also varied per PC and are shown in **Figure 2d**. The highest loading metabolites were 2,3−dihydroxy−5−methylthio−4−pentenoate (DMTPA) for PC1, 5α-androstan-3α,17β-diol monosulfate for PC2, sphingomyelin (d17:1/14:0, d16:1/15:0) for PC3, hydroxy palmitoyl sphingomyelin (d18:1/16:0(OH)) for PC4, and 3-hydroxybutyroylglycine for PC5.

In summary, we reduced the dimensionality of each participant’s metabolome to five principal component scores, which collectively explain 27% of the total population variance and represent specific biochemical profiles.

### Metabolome PCs show distinct functional enrichments

Next, we investigated whether the described metabolome PCs represent concrete biological functions. Metabolite set enrichment analysis (MSEA) on the five sorted lists of metabolome PC loadings revealed that PC2 metabolites with high loadings were significantly enriched for “Transport of small molecules” and “SLC-mediated transmembrane transport” (protein-mediated transfer of molecules between the extra– and intracellular space) while PC3 and PC4 metabolites with high loadings respectively belonged to “Sphingolipid metabolism: integrated pathway” (involved in cell membrane formation, apoptosis and T-cell function) and “Metabolism of amino acids and derivatives“. MSEA revealed no significant results for PC1 or PC5 (**Figure 3a, Suppl. Figure 3a, Suppl. Table 3**). Since MSEA required use of PubChem IDs which were missing in 19-29% of cases per PC, we also used the sub-pathways assigned to each metabolite provided by Metabolon Inc. to retain all metabolites. We found high proportions (> 6%) of metabolites belonging to “Leucine, Isoleucine and Valine metabolism”, “Food component/plant”, and “Androgenic steroids” for PC2, “Sphingomyelins” and “Phosphatidylcholines” for PC3, and “Sphingomyelins”, “Food component/Plant” and “Benzoate metabolism” for PC4. Again, neither PC1 nor PC5 showed strong enrichments in any sub-pathway (**Suppl. Figure 3b**). We finally performed gene ontology analysis (GO) on all Human Metabolome Database (HMDB) documented protein interaction partners of the metabolites per PC. Of note, 13-25% of metabolites could not be used for protein interaction partner retrieval due to missing HMDB IDs (**Suppl. Table 5**). Most PCs showed unique pathways not observed among the top 10 hits for the others (**Figure 3b, Suppl. Table 6**): PC2 metabolite protein interaction partners showed unique enrichments in “Arginine biosynthesis”, “Biosynthesis of amino acids”, “Biosynthesis of cofactors”, “Lysine degradation” and “Pentose and glucuronate interconversions”, while PC3, 4 and 5 showed unique enrichments for “Glutamatergic synapse”, “Glycine, serine and threonine metabolism”/“Porphyrin metabolism” and “Glutathione metabolism”, respectively.

**Figure legend 3.**
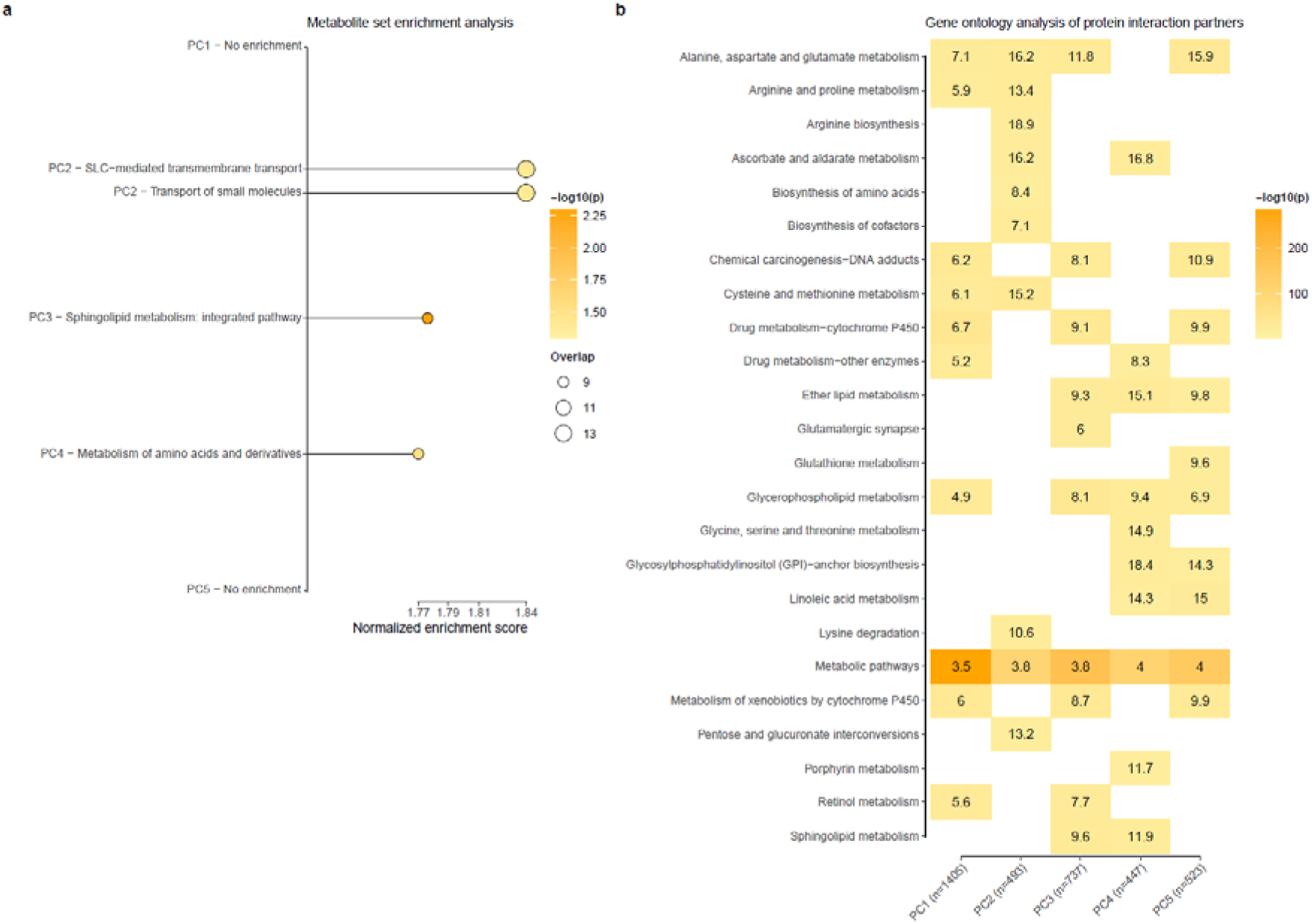
– Functional characterisation of the first five metabolome PCs. **a** Ranking all metabolites per PC by their loading score resulted in five lists used as input for metabolite set enrichment analysis (MSEA) in WebgestaltR. Here, all significant RampDB terms are shown. PC1 and 5 did not show any significant enrichment of metabolite pathways at the top of the ranked input list. The size of the circles refers to the number of overlapping metabolites from the input list and the respective RampDB set. **b** Results from GO analysis of all available HMDB-derived protein interaction partners of significant PC metabolites using ShinyGO. Shown are only the top 10 results per PC. Numbers in the grid signify fold enrichment scores (higher fold enrichment ∼ higher coverage of KEGG set by protein input list). All reported *P*-values have been corrected for multiple testing by the used tools.

Overall, this functional characterisation showed that metabolome profiles can be subdivided into principal components that capture metabolite sets representative of distinct biochemical processes. These components can therefore be used as informative outcomes for further linear modelling to assess determinants of the plasma metabolome.

### Demographics and somatic health have the strongest associations with metabolome PCs, cross-sectionally and longitudinally

Having established the biological significance of metabolomic PC scores, we next aimed to identify baseline determinants of the baseline metabolome and of metabolomic changes over 6 years (**Table 1**). We extracted five metabolome baseline scores for PC1-5 and calculated five metabolome change scores termed ΔPC1-5, which were used as model outcomes.

**Table 1.** Cohort characteristics and determinant metrics.

| Domain | Determinants |  |  | Missing |
| --- | --- | --- | --- | --- |
| Demographic | Sex | Male | 658 (34.6%) | 0 (0%) |
|  |  | Female | 1244 (65.4%) |  |
|  | Age (years) |  | 44 [18, 65] | 0 (0%) |
|  | Education (years) |  | 12 [5, 18] | 0 (0%) |
|  | Degree of urbanisation | Not urbanised | 104 (5.5%) | 0 (0%) |
|  |  | Hardly urbanised | 143 (7.5%) |  |
|  |  | Moderately urbanised | 287 (15.1%) |  |
|  |  | Strongly urbanised | 245 (12.9%) |  |
|  |  | Extremely urbanised | 1123 (59%) |  |
| Psychosocial | Partner | No | 570 (30%) | 0 (0%) |
|  |  | Yes | 1332 (70%) |  |
|  | Work related stress | No work | 430 (22.6%) | 144 (7.6%) |
|  |  | No high strain work | 1121 (58.9%) |  |
|  |  | High strain work | 207 (10.9%) |  |
|  | Leisure activity score |  | 14 [5, 26] | 24 (1.3%) |
|  | Daily hassles score |  | 30 [20, 64] | 19 (1%) |
|  | Childhood trauma score |  | 0 [0, 8] | 4 (0.2%) |
|  | N recent negative life events |  | 0 [0, 7] | 0 (0%) |
| Lifestyle | Alcohol intake (drinks per week) |  | 3.8 [0, 66] | 17 (0.9%) |
|  | Smoking status | Never smoker | 561 (29.5%) | 0 (0%) |
|  |  | Former smoker | 680 (35.8%) |  |
|  |  | Current smoker | 661 (34.8%) |  |
|  | Physical activity (MET-minutes per week) |  | 2856 [0, 19278] | 111 (5.8%) |
| Somatic health | N somatic diseases |  | 1 [0, 7] | 0 (0%) |
|  | N frequent medications |  | 1 [0, 10] | 0 (0%) |
|  | BMI |  | 24.5 [14.7, 55.8] | 1 (0.1%) |
| Mental health | Depression | No diagnosis | 1247 (65.6%) | 0 (0%) |
|  |  | Depression diagnosis | 655 (34.4%) |  |
|  | Anxiety | No diagnosis | 1166 (61.3%) | 0 (0%) |
|  |  | Anxiety diagnosis | 736 (38.7%) |  |

Note: For categorical variables, values are N (%); for continuous variables, values are Median [Min, Max]

First, we assessed whether any particular domain of determinants (demographics, psychosocial environment, lifestyle, somatic health and mental health) explained metabolome PCs or longitudinal changes better than the others. The demographic and somatic health domains explained most variance in PC1 and PC4, respectively (R^2^ = 0.33 and 0.16, **Figure 4a**). The lifestyle domain explained most variance for PC1 (R^2^ = 0.11). For the psychosocial and mental health domain, we observed the highest R^2^ of 0.08 and 0.06 also in PC1. The longitudinal analyses revealed generally weaker associations: We saw that the demographic domain also explained most additional variance for ΔPC1 scores (adj. R^2^ = 0.11 in addition to the base model including only PC-baseline and shipment batch), while the somatic health domain explained most additional variance in ΔPC5 scores (additional adj. R^2^ = 0.08). The lifestyle domain explained most additional variance for ΔPC4 scores (additional adj. R^2^ = 0.01, respectively). For the psychosocial and the mental health domain, the highest additional R^2^ of 0.012 and 0.002 were seen for ΔPC1/3 and ΔPC4, respectively. Overall, the highest explained variance was observed using the full model for PC1 (adj. R^2^ = 0.35) and ΔPC1 (additional adj. R^2^ = 0.12). An overview of all significant determinants per domain model and PC is shown in **Figure 4b**. In brief, all of the 18 chosen determinants showed significant associations across (Δ)PC1-5, with *anxiety diagnosis* being the only exception. An *anxiety diagnosis* showed nominally significant associations in the adjusted mental health models, but it did not withstand Benjamini-Hochberg (BH) correction.

**Figure legend 4.**
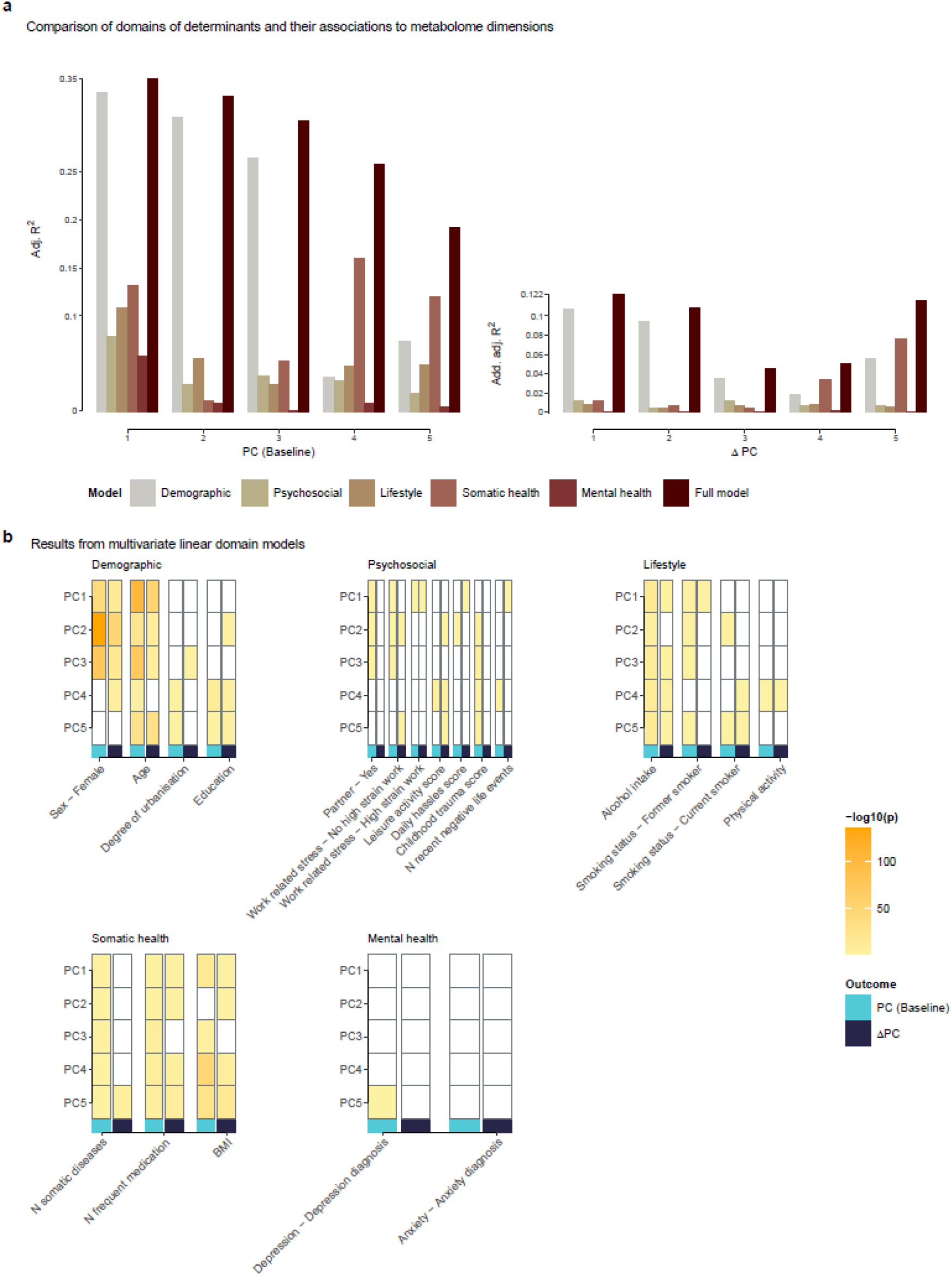
– Linear modelling results from the domain models. **a** Comparison of adjusted R^2^ values from linear regression using the domain specific and full models with baseline determinants, with PC-baseline or ΔPC score as outcomes. **b** Visual overview of significant determinants per domain model. For detailed statistics see ***Suppl. Information***.

As opposed to the separate domain models, the full models provide an overview of the independent associations of all 18 baseline determinants on PC-baseline or ΔPC scores (**Figure 5a,b**). In the following, the individual determinants’ associations with metabolome PCs are listed in order of strong to weak effect sizes. *Sex* emerged as one of the most predictive determinants of metabolome PCs at baseline and for 6-year change, showing associations for PC1-4 and ΔPC1-3. Similarly, *age* was associated with PC1-5 and ΔPC1,2,3,5. The *number of medications* influenced PC2-5 scores and ΔPC2,4,5. *Alcohol intake* was reflected in PC1,3,4,5 and ΔPC4,5. *BMI* showed associations with PC1,4,5 and ΔPC4,5. *Current smoking* was associated with metabolites across PC1-2 and ΔPC5, while *former smoking* showed lasting influences on ΔPC5. *Years of education* were associated with PC4 and ΔPC1,5. *Daily hassles* were associated with PC2 and ΔPC1. *Degree of urbanisation* was linked to PC4,5. *No high strain work* (versus *No work*) and *high strain work* showed associations with ΔPC2. *Leisure activity* and *reported childhood trauma* showed associations with PC4. *Number of somatic diseases* showed an association with ΔPC5. *Partner status*, *physical activity* and a *depression diagnosis* had only nominally significant associations with PC2, ΔPC4,5 and PC5/ΔPC3, respectively, which did not withstand BH correction (**Suppl. Table 7**). An *anxiety diagnosis* and *number of recent negative life events* had no nominally significant associations. All associations between the metabolome PCs, their functional enrichments and significant determinants from the cross-sectional and longitudinal modelling are listed as a summary in **Table 2**. Moreover, a PermANOVA of *age*, *sex*, *smoking status* and *BMI* on n=2803 baseline metabolome measurements revealed that those determinants alone explained 8.9% of variance in our sample (*p* < 0.001).

**Figure legend 5.**
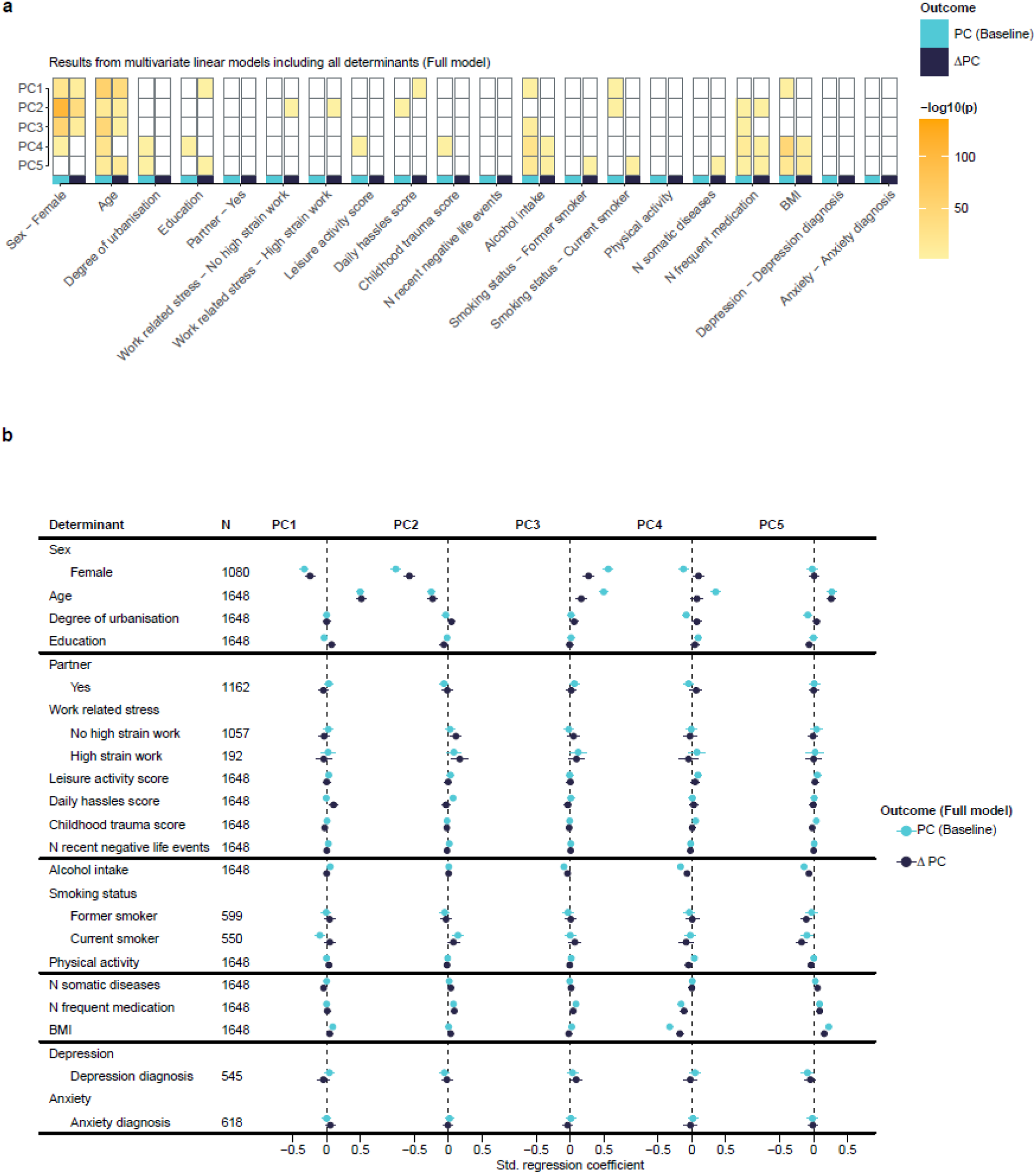
– Linear modelling results from the full models. **a** Visual overview of significant determinants. **b** Forest plot of standardized regression coefficients and 95% confidence intervals for all determinants in the full model. For detailed statistics see ***Suppl. Information***.

**Table 2.** Results summary.

| Metabolome PC | Metabolite class representation | MSEA results | GO results on protein interaction partners<br>(Top 3, ranked by fold enrichment) | Best domain model for PC-baseline and ΔPC | Significant determinants<br>(Full model for PC-baseline score) | Significant determinants<br>(Full model for ΔPC change score) |
| --- | --- | --- | --- | --- | --- | --- |
| PC1<br>(604 metabolites) | General (similar to overall dataset composition) | n.s. | Alanine aspartate and glutamate metabolism<br>Drug metabolism-cytochrome P450<br>Chemical carcinogenesis-DNA adducts | Demographic | Sex, Age<br>Alcohol intake, Current smoking<br>BMI | Sex, Age, Education<br>Daily hassles |
| PC2<br>(257 metabolites) | Less lipids, more amino acids, more xenobiotics, less cofactors and vitamins than PC1 | Transport of small molecules<br>SLC-mediated transmembrane transport | Arginine biosynthesis<br>Alanine aspartate and glutamate metabolism<br>Ascorbate and aldarate metabolism | Demographic | Sex, Age<br>Daily hassles<br>Current smoking<br>Medication | Sex, Age<br>No high strain work, High strain work<br>Medication |
| PC3<br>(232 metabolites) | More lipids, less amino acids, more xenobiotics, less peptides, less nucleotides than PC1 | Sphingolipid metabolism: integrated pathway | Alanine aspartate and glutamate metabolism<br>Sphingolipid metabolism<br>Ether lipid metabolism | Demographic | Sex, Age<br>Alcohol intake<br>Medication | Sex, Age |
| PC4<br>(235 metabolites) | Less amino acids, more xenobiotics, more cofactors and vitamins than PC1 | Metabolism of amino acids and derivatives | GPI-anchor biosynthesis<br>Ascorbate and aldarate metabolism<br>Ether lipid metabolism | Somatic health | Sex, Age, Urbanisation, Education<br>Leisure activity, Childhood trauma<br>Alcohol intake<br>Medication, BMI | Alcohol intake<br>Medication, BMI |
| PC5<br>(180 metabolites) | Less lipids, more amino acids, more peptides, more nucleotides than PC1 | n.s. | Alanine aspartate and glutamate metabolism<br>Linoleic acid metabolism<br>GPI-anchor biosynthesis | Somatic health | Age, Urbanisation<br>Alcohol intake<br>Medication, BMI | Age, Education<br>Alcohol intake, Former smoking, Current smoking<br>Somatic diseases, Medication, BMI |

When we expanded our analysis from domain-specific to full models, we observed several changes. Most notably, for PC2, *daily hassles* remained significantly associated while other psychosocial factors became weaker associated or lost statistical significance. A similar attenuation was noted for *physical activity* and *number of recent negative life events* and all PC-baseline associations for *number of somatic diseases*.

Given the high influence of age, sex and BMI on metabolite levels as seen in the analysis above, additional domain model results adjusted individually for age, sex and BMI can be found in **Suppl. Table 7**. In general, subsequent results were highly similar to the full model results, indicating that the associations between certain determinants and metabolite PCs were attenuated by these important determinants.

Taken together, these results showed that baseline determinants explained a larger proportion of variance in PC-baseline scores as compared to ΔPC scores and that metabolome PC1-5 are differentially associated with the five domains. Across most metabolome PCs, the R^2^ increased predominantly by adding information on somatic health and lifestyle beyond demographic characteristics. We further observed that, while the main influences on the metabolome in our study pertained to sex, age, alcohol intake, BMI and medication, a wide variety of other factors contributed additional but subtler influences. These subtle influences were more abundant in domain models, when fewer mediators and confounders were present. Lastly, we observed that determinants linked to the baseline metabolome profile did not necessarily have significant associations with changes over 6 years and vice versa.

## Discussion

In this analysis of the plasma metabolome, we examined five metabolome PCs derived from principal component analysis and their associations with participant characteristics across demographic, psychosocial, lifestyle, somatic and mental health domains. Principal component, metabolite set enrichment and Metabolon Inc. sub-pathway analysis revealed that metabolome PCs captured distinct biological processes. Most importantly, gene ontology analysis on metabolites’ protein interaction partners reflected potential influences of specific exposures: PC2, primarily influenced by demographic factors (especially sex), showed enrichment in pentose and glucuronate interconversions linked to Uridine 5’-diphospho-glucuronosyltransferase (UGT) enzymes which mediate the inactivation of e.g. oestrogens via glucuronidation. In the domain model analysis, it was PC2 which had the most significant associations with psychosocial factors out of all five PCs, next to PC1. PC4, predominantly shaped by somatic health factors including medication use and especially BMI, was enriched for drug metabolism and porphyrin metabolism, an essential pathway for haemoglobin synthesis, and glycine-serine-and-threonine metabolism, a pathway which has been observed for maternal obesity in cord blood and general adiposity (19, 20). PC5 showed consistent associations with lifestyle factors and medication and was uniquely enriched for glutathione metabolism, reflecting the antioxidant and detoxification demands associated with alcohol consumption, smoking and medication use (21).

We observed that our chosen determinants explained a limited amount of additional variance in metabolite change over time beyond that already captured by baseline metabolite levels. This is likely due to the restricted range of variability in 6-year change in our longitudinal analysis, which is consistent with previous studies, showing similar metabolomic stability over several years of follow-up (4, 22, 23). Strikingly, *age* influenced all metabolome change scores except for ΔPC4. Other studies also found that age had a strong influence on longitudinal metabolomic trajectories (24, 25). In fact, the robust association between *age* and the metabolome likely explains why *number of somatic diseases* and *number of recent negative life events*, both correlated with age though with different directionality, showed fewer significant independent associations in the full model. Notably, many other determinants in our study showed significant associations with longitudinal metabolomic change in our analysis, but these did not seem to follow a systematic pattern. *Education* is worth highlighting in this context, as it showed significant longitudinal associations with ΔPC5 in both the domain and full model. Due to its close link to socioeconomic status, this underlines the long-term potential of socioeconomic inequality to influence the metabolism (26, 27). When focusing on individual metabolites, we observed strong reductions in PFAS molecules over 6 years, potentially due to legislative efforts aimed at reducing the industrial use of these synthetic chemicals. Moreover, the neurosteroids dehydroepiandrosterone sulfate (DHEA-S) and the closely related pregnenolone sulfate (PS) were also among the most decreasing metabolites over 6 years. DHEA-S and PS have been investigated for their memory-enhancing and antidepressant effects and were shown to attenuate memory deficits in Alzheimer’s disease induced by beta-amyloid β25-35 (28, 29). They have also been discussed as potential therapeutics for several psychiatric conditions (30). Therefore, the observed reduction in DHEA-S and PS over 6 years in our cohort could be linked to detrimental aging phenotypes, although a prominent study has also revealed no beneficial effects of DHEA supplementation (31). Among the highest increasing compounds, we found argininate, dibutyl sulfosuccinate and sarcosine. The arginine metabolism is shown to be involved in cancer malignancy (32). Dibutyl (sodium) sulfosuccinate is a synthetic surfactant used in pharmaceuticals, detergents as well as personal care products (33). Contrastingly, sarcosine has previously been found to decrease with age in mice (34). Importantly, these results stem from a paired, but unadjusted comparison of baseline and FU6 data including all 1902 participants and might therefore differ if important covariates such as age, sex, BMI, smoking, medication use or health status are included.

Our findings further revealed that different metabolome PCs are differentially shaped by the assessed domains, reflecting the notion that exposures might leave specific rather than broad traces in the organism. The most pronounced effects were observable for *sex*, *age* (demographics), *medication*, *BMI* (somatic health), and lastly *alcohol consumption* and *smoking* (lifestyle). These associations align with previous studies and likely occur via interaction with established biological mechanisms (3–5), including the influence of sex hormones (35), aging and mortality-related metabolomic processes (25, 36, 37), microbial metabolite production (38–40) and xenobiotics exposure (41) which collectively shape the level of circulating metabolites. Our cross-sectional PermANOVA R^2^ estimate of 8.9% for the combined effects of *age*, *sex*, *BMI*, and *smoking* on the metabolome exceeds previous reports by almost double, although this may partly be explained by differences in metabolite coverage and technical factors (4). Interestingly, *former smoking* demonstrated weaker associations with metabolome PCs than *current smoking*, suggesting the possibility of metabolic recovery following smoking cessation. Furthermore, *leisure activity* had one significant association with PC4 even after adjusting for *alcohol intake*, *smoking*, *physical activity*, and *partner status*. This suggests an influence of spending free time outdoors, a finding that warrants structured follow-up analyses. While *leisure activity* and *physical activity* share some overlapping elements, leisure activity primarily reflects social activity, whereas physical activity mainly captures sports, exercise and movement. The consistent association of *degree of urbanisation* with PC4,5 in the demographic domain model and in the full model suggests that exposure to city environments might lead to a metabolic signature presumably shaped by airborne xenobiotics and toxins, but also lifestyle factors which differ between the rural and the urban population. Similar observations in plasma have been made by a recent Chinese study (42).

Psychological stress emerged as another contributor to metabolome profiles, with correlations observed across multiple stressors including *daily hassles*, *occupational stress*, and *childhood trauma*. These stress-related traces, while more subtle than those from other domains, may occur through multiple pathways including epigenetic modifications (43–46), altered microbial metabolite production (47), or behavioural adaptations affecting body composition and sleep patterns (48, 49). Influences of stress on the metabolome have been studied previously but none explicitly considered non-pathological occupational or daily stress in a large, representative cohort (12). Our findings on childhood trauma and its metabolomic signature recapitulate previous work from NESDA, which showed a dose-response relationship between childhood trauma and specific plasma metabolites (17). In line with previous studies (8, 50, 51) a depressive disorder diagnosis (MDD and/or dysthymia) was associated in the mental health domain model with metabolites captured in our study by PC5. This association was reduced and no longer statistically significant in the full model including all determinants, which may suggest that the association between depression and metabolite levels could be largely explained by relevant factors included in the full model. Nevertheless, it is important to remark that estimates from the full model should be interpreted with caution due to potential overadjustment, as for each determinant all other factors may represent not only confounders to be taken into account but also mediators (e.g. unhealthy lifestyle as a consequence of depression) or colliders (e.g. chronic diseases determined by both depression and metabolic alterations) that should not be included in the model for proper interpretation.

This study represents the first large-scale investigation to simultaneously examine demographic, psychosocial, lifestyle, somatic and mental health determinants of the metabolome using both cross-sectional and longitudinal data. The comprehensive nature of the chosen determinants enables direct comparison of explained variance across domains and provides a basis for more targeted mechanistic investigations. However, several limitations warrant consideration. The reliance on PCA, while facilitating dimensional interpretation, necessarily involves loss of information compared to analyses on a metabolite-level. Additionally, HMDB protein interaction partners were used for gene ontology analysis, which also includes connections between proteins and metabolites that have not been validated. Furthermore, the ethnic, genetic and geographic homogeneity of our Dutch cohort may limit generalizability to other populations. Finally, untargeted metabolomic analyses are highly sensitive to laboratory protocols, which can introduce noise through anti-clotting agents, sample storage, shipment, and handling procedures (52). However, the large sample size and inclusion of shipment as a covariate should be sufficient to mitigate technical confounders in this study.

Future research should focus on disentangling the overlapping influences of the here presented determinants on the metabolome, which will help to better understand the mechanisms that can either harm or protect metabolic health. Moreover, follow-up investigations should zoom in on individual metabolites to identify specific biomarkers underlying the associations presented here, especially exposure to medication/antidepressants, which usually have distinct metabolic effects which are hard to capture with principal components. Given its distinct metabolite class composition, PC2 metabolites emerged as particularly interesting due to their characteristic pathway enrichment, strong associations with sex and age and associations with psychosocial factors (including psychological stress) in the domain and full models, respectively. Comparative analysis of PC2 metabolites between both sexes could enhance our understanding of the molecular mechanisms underlying psychological stress exposures and inform targeted interventions with potential relevance for females, who are at higher risk of developing stress-related diseases such as depression (53, 54).

In conclusion, this study represents the largest epidemiological investigation to date examining a wide variety of determinants that influence metabolite levels in plasma cross-sectionally and longitudinally. Our approach extends previous research by investigating metabolome PCs rather than the metabolome as a whole, and by examining not only standard factors but also associations with psychosocial and lifestyle factors. Through a comprehensive analysis across five conceptual domains, we discovered that many previously unknown determinants beyond demographics shape the plasma metabolome. These findings open up new questions of how life exposures are embedded into the metabolome and how these could inform therapeutic interventions or prevention options for conditions involving metabolic dysregulation.

## Methods

### Study design and participants

Data are from the Netherlands Study of Depression and Anxiety (NESDA), an ongoing multi-centre, observational, naturalistic and longitudinal cohort study examining the course and consequences of depressive and anxiety disorders. A detailed description of the study rationale, design and methods is given elsewhere (55). Briefly, 2981 adults (18-65 years old) with or without a diagnosis of depression and/or anxiety disorder were recruited from community samples, primary care practices, and mental health organisations between 2004 and 2007. Detailed assessments were repeated after one, two, four and 6 years of follow-up. Exclusion criteria were being diagnosed with a current clinically overt psychiatric disorder other than depression and anxiety as well as lacking proficiency in Dutch. Ethical approval was obtained centrally from the Ethical Review Board of the Vrije Universiteit Medical Centre (reference number 2003/183) in Amsterdam, the Netherlands, as well as from the Ethics Review Boards of the other participating research centres. All participants provided written informed consent.

### Metabolomic data generation and preprocessing

Plasma metabolite measurement and subsequent data pre-processing have been described elsewhere extensively (8). Data was analysed and pre-processed in line with protocols adopted by established international consortia (https://www.omicscience.org/), log_2_-transformed and winsorised. In brief, fasting EDTA plasma samples were collected in the morning at baseline and FU6 and were stored at –80°C. Samples were sent for analysis where metabolome profiles were assessed using the mass spectrometry-based untargeted HD4 platform from Metabolon Inc. (Durham, NC, USA), with paired samples being processed on the same plate within the same batch. Shipment was performed in two batches. In total, 4827 samples were measured, 2812 from baseline and 2015 from FU6. From that, 1902 study participants had available data across the two timepoints. From 820 measured metabolites in total, 139 unknown metabolites and the anti-clotting agent EDTA were excluded. Thus, 680 metabolites were available for analysis. Mutually exclusive labels provided by Metabolon Inc. were used to categorise metabolite classes: lipids, amino acids, xenobiotics, peptides, cofactors and vitamins, nucleotides, carbohydrates, partially characterised molecules and energy.

### Clinical assessments and other measures at baseline

In total, 18 exposome factors (determinants) from five conceptual domains were selected based on their known general relevance for human metabolism, physiology and/or their influence on stress levels (42, 56–66). Of note, we did not observe concerning levels of multicollinearity across our chosen determinants (max. Pearson’s *r* = 0.48 for N somatic diseases ∼ N frequent medication), **Suppl. Figure 4**).

Demographic: *Sex*, *age* and *years of education* were assessed via basic intake questionnaires at study begin. The *degree of urbanisation* was determined based on current zip code and population density information from the Central Bureau of Statistics of the Netherlands.

Psychosocial environment: *Partner status* was assessed via a basic intake questionnaire at study begin. *Work related stress* was assessed using the Karasek questionnaire (67), with high strain work being defined as current work with self-reported high job demand scores (≥0.5) and low job control scores (≤0.5). Job demand refers to the expected pace of work, workload and work intensity whereas job control assesses self-perceived autonomy at the workplace. The *leisure activity score* is a summed score of five items from a questionnaire asking about the current frequency of social free time activities such as going to a bar, museum, restaurant, association, amusement park, cinema, club or sports activity outside of home. The *daily hassles score* was derived using a questionnaire asking about daily life social stressors such as interpersonal difficulties or conflicts with friends, colleagues or family, high mental load, rejection, disappointment or financial struggles in the past month. The reported *childhood trauma score* was assessed using the Childhood Trauma Questionnaire from the NEMESIS study (68) and asked about traumatic experiences such as parental divorce, juvenile prison detention, foster family or child home placement, emotional neglect and psychological, physical or sexual abuse in the first 16 years of life. The *number of negative recent life events* was determined based on an abridged version of the Brugha questionnaire (69) which asks about the occurrence of adverse events in the past year, such as death or serious illness of loved ones, job loss, terminated relationship or friendship, contact with the police or court and other events.

Lifestyle: *Alcohol intake* was calculated based on data originating from the AUDIT Questionnaire and is expressed as drinks per week (70). *Smoking status* was determined based on data originating from the Fagerström Questionnaire and is expressed as never smoker, former smoker and current smoker (71). *Physical activity* was assessed using the International Physical Activity Questionnaire and is expressed as MET-minutes per week (72), capturing sports activity, movement and commuting.

Somatic health: We deduced the *number of somatic diseases* of participants based on a questionnaire asking about the following current, chronic conditions: asthma, heart disease, diabetes, stroke, arthritis, cancer, hypertension, intestinal disorders, liver disease, epilepsy, chronic fatigue, allergies, thyroid gland disease, injury in past year, head injury (ever), and others. *Number of medications* was defined as drugs taken on ≥50% of the days in a current month for conditions falling under a WHO ATC classification. Dietary supplements and homeopathic medication were excluded. The top 10 most frequently used medications were paracetamol, ibuprofen, paroxetine, oxazepam, omeprazole, metoprolol, citalopram, venlafaxine, amiloride and simvastatin. For the frequency of ATC codes in our sample see **Suppl. Table 8**. *BMI* was determined as current body weight [kg] /height^2^ [m].

Mental health: The “*depression diagnosis*” variable in this study encompasses CIDI diagnosed major depressive disorder or dysthymia in the past 6 months. The “*anxiety diagnosis*” encompasses CIDI diagnosed generalised anxiety disorder, agoraphobia, panic disorder or social phobia in the past 6 months.

### Statistical analyses

If not stated otherwise, all analyses were conducted in R (version 4.3.3) in Microsoft VS Code. See **Suppl. Information** for further details on packages used for analysis and plotting (excl. dependencies).

Intraclass correlation: We assessed intraclass correlations between baseline and FU6 for 680 metabolites to assess within-subject change using the ICC function from the R psych package. We reported ICC1.

Principal component analysis: We performed principal component analysis on a data matrix of 3804 samples from baseline and FU6 x 680 metabolites features using the prcomp function from base R. The scale and center arguments were set to TRUE. We calculated a metabolome change score per PC for each sample as ΔPC = PC_FU6_ – PC_Baseline_. To perform a significance test for principal components themselves and the respective loadings for each principal component, we used the PCAtest function from the PCAtest R package (73). All PCs were significant, while only a subset of metabolites represented significant loadings for each PC. Significance was determined based on 100 permutations x 100 bootstraps of the original data matrix. For internal validation of the PCA results (scree plot and loadings) this was repeated for 2812 – 1902 = 910 additional baseline samples, including NESDA study participants and sibling samples.

Metabolite ids: For each metabolite, Metabolon Inc. provided the chemical name, its PubChem ID and its HMDB ID (74, 75). PubChem IDs had lower missingness than HMDB IDs and were thus used for metabolite set enrichment analysis (MSEA). For gene ontology analyses (GO), protein interaction partners of metabolites per PC1-5 were used as input and expressed as gene symbols. The HMDB API was used to derive those protein interaction partners, requiring HMDB IDs as queries. Therefore, different IDs were used compared to MSEA. In cases where multiple IDs were available per metabolite as provided by Metabolon Inc., the first one listed was chosen.

Metabolite set enrichment analysis: To identify enrichments of certain functional pathways along a metabolite list (ranked by metabolite loading score per PC from highest to lowest), we used the WebGestaltR function of the WebGestaltR package, setting enrichMethod to “GSEA“, organism to “hsapiens“, enrichDatabase to “pathway_RampDB_Metabolomics “, interestGeneType to “pubchem” and otherwise standard settings (76). All RampDB metabolites were considered as background. Higher normalised enrichment scores indicate enrichment of metabolites closer to the top of the list, where the most important metabolites for the respective PC are found.

Gene ontology analysis: To identify significant overlaps between metabolite protein interaction partners per PC and Kyoto Encyclopaedia of Genes and Genomes (KEGG) pathway sets, we used the ShinyGO v0.82 website https://bioinformatics.sdstate.edu/go/ (77) with “Use pathway DB for gene counts” set to TRUE, showing the top 10 enriched pathways with standard settings. KEGG was chosen to investigate general human biochemical pathways (78).

Linear regression: To model PC-baseline and ΔPC scores (outcomes) on baseline determinants (predictors), we used the lm function from base R. Shipment batch was included as covariate for both outcomes, the respective PC-baseline score was included as a covariate for all ΔPC models. Outcomes and all numerical predictors were scaled (“robust scaling” method, using the ratio of median and interquartile range). Degree of urbanisation was an ordinal variable. Smoking status and work-related stress were categorical variables. Sex, partner status, depression diagnosis and anxiety diagnosis were dichotomous variables. All other variables were numeric. To compare the models, R^2^ coefficients of determination were adjusted for the number of predictors in the model. Sample sizes per model differed depending on missingness of data for some determinants. Sensitivity analyses restricted to participants with complete data (n=1648) yielded nearly identical results (correlations with original adj. R²: Pearson’s *r* = 0.999 for PC-baseline models, *r* = 0.995 for ΔPC models), confirming our findings’ robustness. Nominal *P*-values were corrected using the BH method per linear model (79).

As part of the longitudinal analysis, we made use of a base model in addition to the five domain models, which included only the respective PC-baseline score and the shipment covariate. This ensures that we quantify the variance of metabolome change scores solely explained by baseline metabolome PCs and technical confounding to understand the additional explained variance coming from the determinants of interest in the other models. Therefore, “additional explained variance” refers to adj. R^2^ from which the base model adj. R^2^ has been subtracted.

PermANOVA: To model the whole metabolite matrix on a set of determinants for comparison purposes to a previous publication, we used the adonis2 function from the vegan R package with 1000 permutations and the “euclidean” method. Since only baseline metabolomics data was assessed, we included samples without follow-up measurement, increasing sample size to n=2803 study participants of whom the respective data was collected.

*(For methods used for Suppl. Figures see **Suppl. Information**)*

## Funding declaration

The infrastructure for the NESDA study (https://www.nesda.nl/) is funded through the Geestkracht program of the Netherlands Organisation for Health Research and Development (ZonMw, grant number 10-000-1002) and financial contributions by participating universities and mental health care organizations (VU University Medical Center, GGZ inGeest, Leiden University Medical Center, Leiden University, GGZ Rivierduinen, University Medical Center Groningen, University of Groningen, Lentis, GGZ Friesland, GGZ Drenthe, Rob Giel Onderzoekscentrum). This work was supported by the National Institute of Mental Health (Grant No. <u>R01MH108348</u>) and by a series of grants issued through National Institute on Aging (Grant Nos. <u>U19AG063744</u> and <u>U01AG061359</u> [to principal investigator, RKD]), which supported a large number of scientists working on metabolomics, neuropsychiatric disorders and acylcarnitines. RKD has also received funding from the NIA (Grant No. 1RF1AG058942, 1RF1AG057452, RF1AG051550, U01AG088562, RF1AG059093, and R01AG046171) and FNIH: #DAOU16AMPA. This funding enabled consortia that she leads including the Mood Disorder Precision Medicine Consortium (MDPMC), the Alzheimer’s Disease Metabolomics Consortium (ADMC), and the Alzheimer Gut Microbiome Project (AGMP) that contributed to the metabolomic analyses of the NESDA sample used in this article and to acylcarnitine discoveries. A listing of AGMP Investigators can be found at https://alzheimergut.org/meet-the-team/. A listing of ADMC investigators can be found at: https://sites.duke.edu/adnimetab/team/. A listing of MDPMC investigators can be found at https://sites.duke.edu/mdpmc/files/2022/06/MDPMC-Members2022.pdf. The funders listed above had no role in the design or conduct of the study; the collection, management, analysis, or interpretation of the data; preparation, review, or approval of the article; or the decision to submit the article for publication. In addition, DK and BWJHP are supported by the research project <u>Stress in Action – Innovation in the understanding and application of daily-life stress.</u> which is financially supported by the Dutch Research Council and the Dutch Ministry of Education, Culture and Science (NWO gravitation Grant No. <u>024.005.010</u>). YM has received funding from Amsterdam UMC (Starter Grant Ronde 2), Amsterdam Neuroscience (PoC funding 2024–2026) and <u>ImmunoMIND</u>, funded by UK Research and Innovation as part of the UK national Mental Health Platform. LKMH was funded by the Veni award (Grant No. 09150162210201) from the Dutch Research Council (NWO).

## Conflict of interest

Dr. Kaddurah-Daouk is an inventor on a series of patents on use of metabolomics for the diagnosis and treatment of central nervous system diseases and holds equity in Metabolon Inc., Chymia, and Metabosensor. Drs. Arnold and Kastenmüller are inventors of patents on applications of metabolomics in neurodegenerative/neuropsychiatric diseases and hold equity in Chymia LLC, which had no role in this work. The other authors declare no conflict of interest.

## Acknowledgements

We thank all participants and their families for their time, data, and samples. Additionally, we thank the Amsterdam Psychiatry Department’s HPC for technical support during the computational analyses and Rick Jansen for metabolomic data management. Generative AI (Anthropic Claude Sonnet 3.7/4.5) has been used to support coding.

## Author contributions

DK: Conceptualization, Methodology, Formal analysis, Writing – Original Draft, Visualization; YM: Conceptualisation, Methodology, Writing – Review & Editing, Supervision; LKMH: Conceptualisation, Methodology, Writing – Review & Editing, Supervision; NJAvdW: Writing – Review & Editing; MA: Writing – Review & Editing; CRB: Writing – Review & Editing; SM: Writing – Review & Editing; GK: Data generation, Writing – Review & Editing; RKD: Data generation, Writing – Review & Editing; BWJHP: Data collection and generation, Conceptualisation, Writing – Review & Editing, Supervision

## Data and code availability

Code is available from Zenodo at https://doi.org/10.5281/zenodo.17869122.

The data used to support the findings of this study are available upon reasonable request from NESDA, Amsterdam. Information on how to request the study data, including the data sharing policy, can be found at https://www.nesda.nl/nesda-english/.

## Notes

### Funding Statement

The infrastructure for the NESDA study is funded through the Geestkracht program of the Netherlands Organisation for Health Research and Development (ZonMw, grant number 100001002) and financial contributions by participating universities and mental health care organizations (VU University Medical Center, GGZ inGeest, Leiden University Medical Center, Leiden University, GGZ Rivierduinen, University Medical Center Groningen, University of Groningen, Lentis, GGZ Friesland, GGZ Drenthe, Rob Giel Onderzoekscentrum). This work was supported by the National Institute of Mental Health (Grant No. R01MH108348) and by a series of grants issued through National Institute on Aging (Grant Nos. U19AG063744 and U01AG061359 [to principal investigator, RKD]), which supported a large number of scientists working on metabolomics, neuropsychiatric disorders and acylcarnitines. RKD has also received funding from the NIA (Grant No. 1RF1AG058942, 1RF1AG057452, RF1AG051550, U01AG088562, RF1AG059093, and R01AG046171) and FNIH: DAOU16AMPA. This funding enabled consortia that she leads including the Mood Disorder Precision Medicine Consortium (MDPMC), the Alzheimer's Disease Metabolomics Consortium (ADMC), and the Alzheimer Gut Microbiome Project (AGMP) that contributed to the metabolomic analyses of the NESDA sample used in this article and to acylcarnitine discoveries. The funders listed above had no role in the design or conduct of the study; the collection, management, analysis, or interpretation of the data; preparation, review, or approval of the article; or the decision to submit the article for publication. In addition, DK and BWJHP are supported by the research project Stress in Action, Innovation in the understanding and application of daily-life stress. which is financially supported by the Dutch Research Council and the Dutch Ministry of Education, Culture and Science (NWO gravitation Grant No. 024.005.010). YM has received funding from Amsterdam UMC (Starter Grant Ronde 2), Amsterdam Neuroscience (PoC funding 2024-2026) and ImmunoMIND, funded by UK Research and Innovation as part of the UK national Mental Health Platform. LKMH was funded by the Veni award (Grant No. 09150162210201) from the Dutch Research Council (NWO).

### Author Declarations

Ethical approval was obtained centrally from the Ethical Review Board of the Vrije Universiteit Medical Centre (reference number 2003/183) in Amsterdam, the Netherlands, as well as from the Ethics Review Boards of the other participating research centres. All participants provided written informed consent.

### Summary of Updates

Corrected statistical term in the abstract

