## Supplementary Figures for "Determinants of the plasma metabolome: cross-sectional and longitudinal associations over 6 years in the NESDA cohort"

### Slide 1
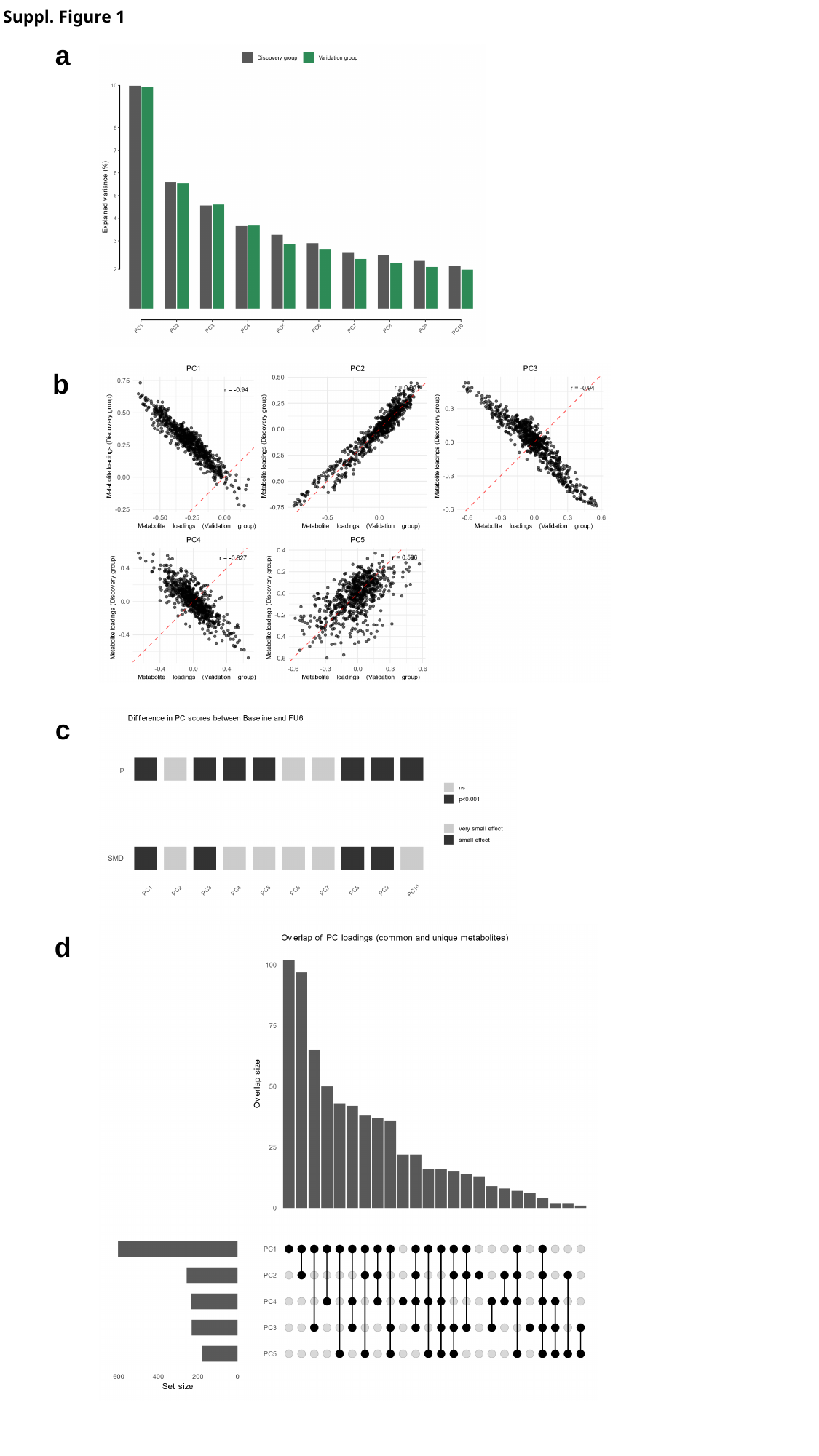

Suppl. Figure 1
a
b
c
d

### Slide 2
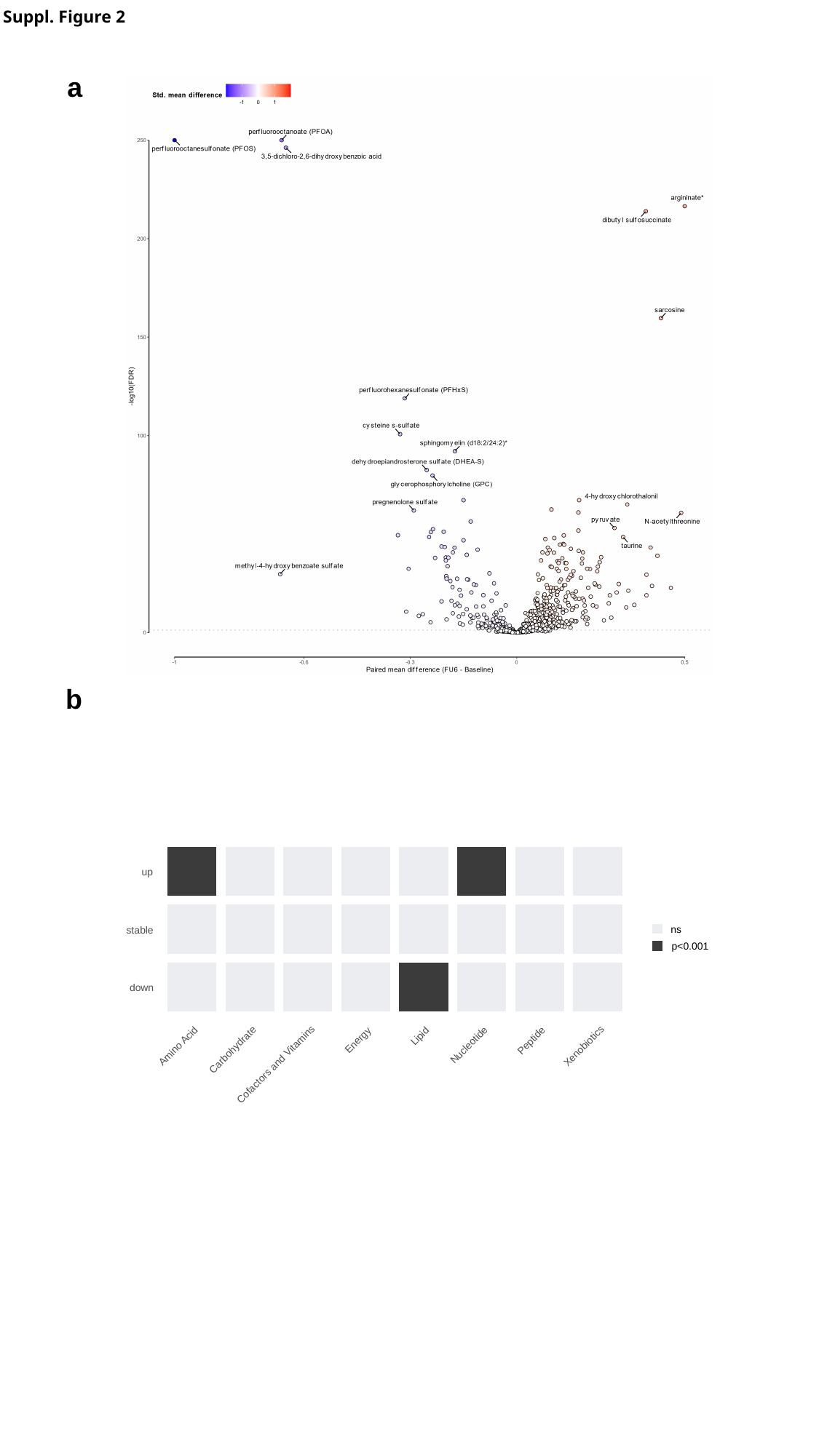

Suppl. Figure 2
a
b
up
ns
stable
p<0.001
down
Lipid
Energy
Peptide
Nucleotide
Amino Acid
Xenobiotics
Carbohydrate
Cofactors and Vitamins

### Slide 3
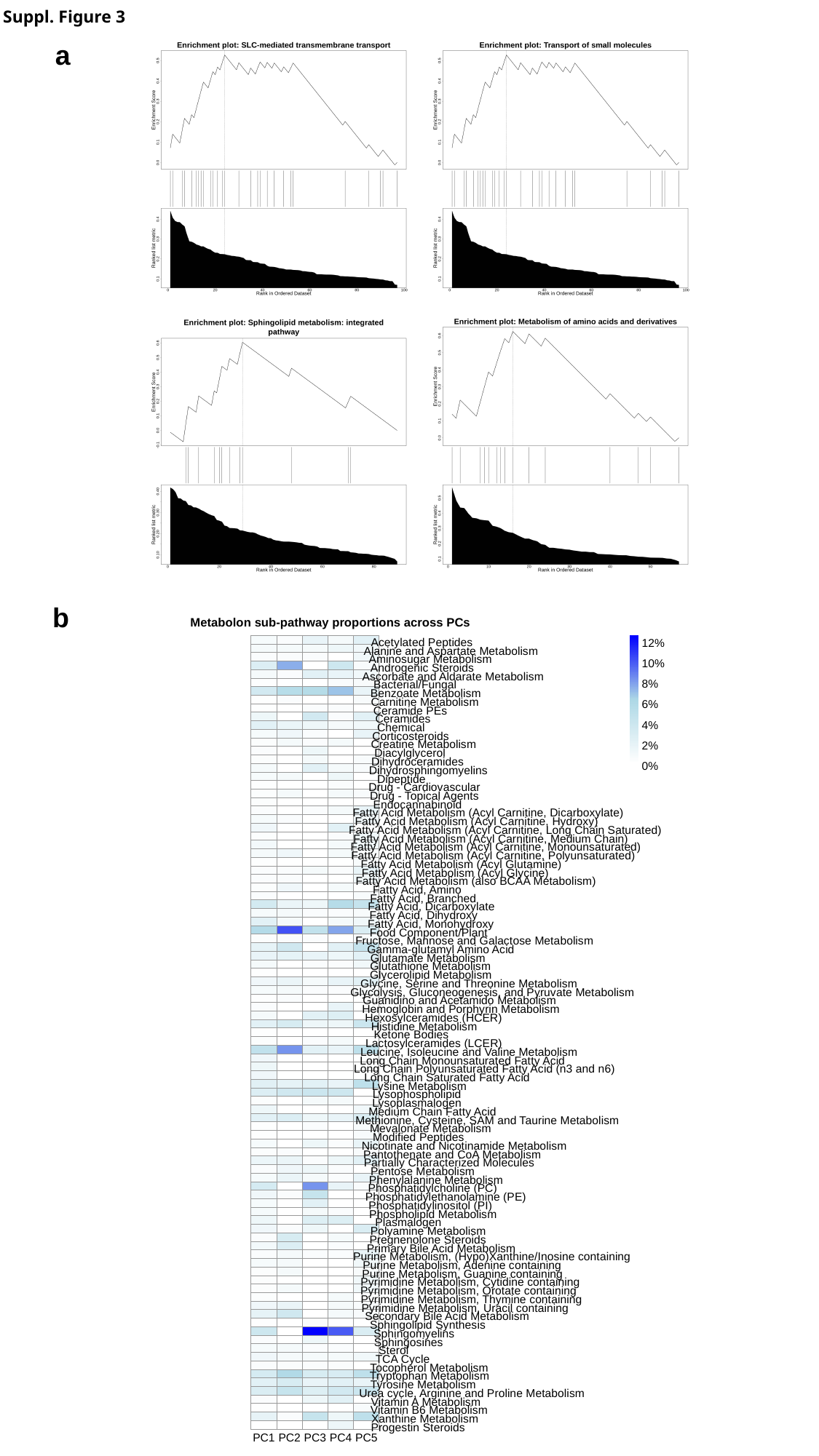

Suppl. Figure 3
a
b
Metabolon sub-pathway proportions across PCs
Acetylated Peptides
12%
Alanine and Aspartate Metabolism
Aminosugar Metabolism
10%
Androgenic Steroids
Ascorbate and Aldarate Metabolism
8%
Bacterial/Fungal
Benzoate Metabolism
Carnitine Metabolism
6%
Ceramide PEs
Ceramides
4%
Chemical
Corticosteroids
Creatine Metabolism
2%
Diacylglycerol
Dihydroceramides
0%
Dihydrosphingomyelins
Dipeptide
Drug - Cardiovascular
Drug - Topical Agents
Endocannabinoid
Fatty Acid Metabolism (Acyl Carnitine, Dicarboxylate)
Fatty Acid Metabolism (Acyl Carnitine, Hydroxy)
Fatty Acid Metabolism (Acyl Carnitine, Long Chain Saturated)
Fatty Acid Metabolism (Acyl Carnitine, Medium Chain)
Fatty Acid Metabolism (Acyl Carnitine, Monounsaturated)
Fatty Acid Metabolism (Acyl Carnitine, Polyunsaturated)
Fatty Acid Metabolism (Acyl Glutamine)
Fatty Acid Metabolism (Acyl Glycine)
Fatty Acid Metabolism (also BCAA Metabolism)
Fatty Acid, Amino
Fatty Acid, Branched
Fatty Acid, Dicarboxylate
Fatty Acid, Dihydroxy
Fatty Acid, Monohydroxy
Food Component/Plant
Fructose, Mannose and Galactose Metabolism
Gamma-glutamyl Amino Acid
Glutamate Metabolism
Glutathione Metabolism
Glycerolipid Metabolism
Glycine, Serine and Threonine Metabolism
Glycolysis, Gluconeogenesis, and Pyruvate Metabolism
Guanidino and Acetamido Metabolism
Hemoglobin and Porphyrin Metabolism
Hexosylceramides (HCER)
Histidine Metabolism
Ketone Bodies
Lactosylceramides (LCER)
Leucine, Isoleucine and Valine Metabolism
Long Chain Monounsaturated Fatty Acid
Long Chain Polyunsaturated Fatty Acid (n3 and n6)
Long Chain Saturated Fatty Acid
Lysine Metabolism
Lysophospholipid
Lysoplasmalogen
Medium Chain Fatty Acid
Methionine, Cysteine, SAM and Taurine Metabolism
Mevalonate Metabolism
Modified Peptides
Nicotinate and Nicotinamide Metabolism
Pantothenate and CoA Metabolism
Partially Characterized Molecules
Pentose Metabolism
Phenylalanine Metabolism
Phosphatidylcholine (PC)
Phosphatidylethanolamine (PE)
Phosphatidylinositol (PI)
Phospholipid Metabolism
Plasmalogen
Polyamine Metabolism
Pregnenolone Steroids
Primary Bile Acid Metabolism
Purine Metabolism, (Hypo)Xanthine/Inosine containing
Purine Metabolism, Adenine containing
Purine Metabolism, Guanine containing
Pyrimidine Metabolism, Cytidine containing
Pyrimidine Metabolism, Orotate containing
Pyrimidine Metabolism, Thymine containing
Pyrimidine Metabolism, Uracil containing
Secondary Bile Acid Metabolism
Sphingolipid Synthesis
Sphingomyelins
Sphingosines
Sterol
TCA Cycle
Tocopherol Metabolism
Tryptophan Metabolism
Tyrosine Metabolism
Urea cycle, Arginine and Proline Metabolism
Vitamin A Metabolism
Vitamin B6 Metabolism
Xanthine Metabolism
Progestin Steroids
PC1
PC2
PC3
PC4
PC5

### Slide 4
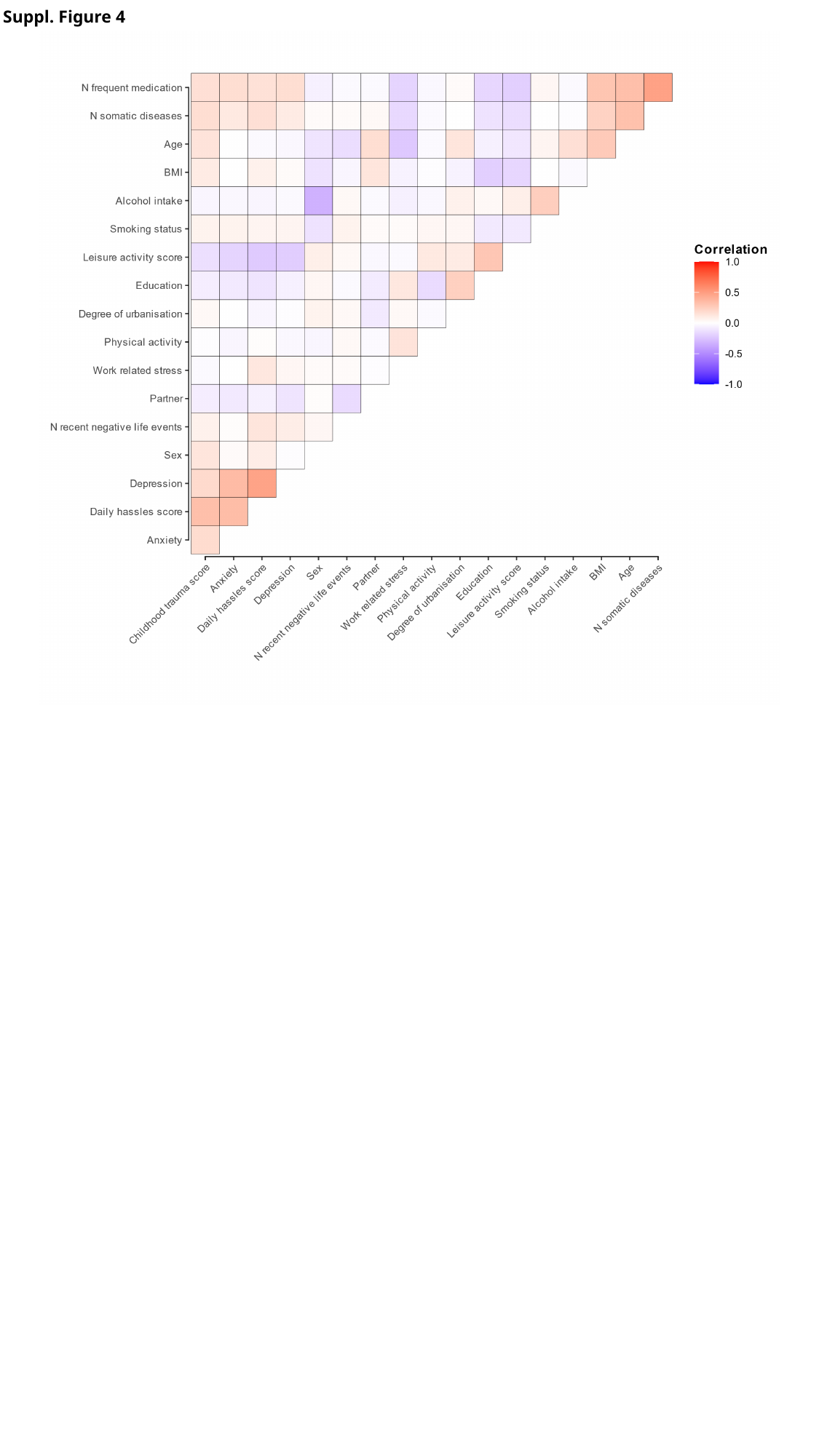

Suppl. Figure 4
