## Supplementary Information for "Determinants of the plasma metabolome: cross-sectional and longitudinal associations over 6 years in the NESDA cohort"

**Suppl. Information**

*Statistical analyses*

Used R packages: Cairo (v1.6-2), ComplexUpset (v1.3.3), corrplot (v0.95), devtools (v2.4.5), dplyr (v1.1.4), flextable (v0.9.7), forcats (v1.0.0), forestploter (v1.1.3), ggplot2 (v3.5.1), ggplotify (v0.1.2), ggpubr (v0.6.0), ggrepel (v0.9.6), ggsignif (v0.6.4), ggthemes (v5.1.0), gridExtra (v2.3.0), haven (v2.5.4), htmltools (v0.5.8.1), httpgd (v2.0.3), httr (v1.4.7), jsonlite (v2.0.0), khroma (v1.16.0), labelled (v2.14.0), officer (v0.6.7), openxlsx (v4.2.8), pandoc (v0.2.0), patchwork (v1.3.0), PCAtest (v0.0.2), polycor (v0.8-1), psych (v2.5.6), readr (v2.1.5), readxl (v1.4.3), rstatix (v0.7.2), scales (v1.3.0), stringr (v1.5.1), tibble (v3.2.1), tidyr (v1.3.1), tidyverse (v2.0.0), WebGestaltR (v1.0.0), vegan (2.7-1).

Intrapersonal changes of individual metabolites: The volcano plot in supplementary material was generated with results from repeated paired two-sample t-tests for baseline vs FU6 to assess changes of individual metabolites within the same person. *P*-values were corrected for multiple testing using the BH-method. Metabolites not showing statistically significant differences between assessments were deemed stable, whereas significant hits were considered increasing or decreasing based on the positive or negative sign of the paired mean difference (FU6 - baseline), respectively. Standardised mean differences (SMD) were calculated as mean paired differences divided by the standard deviation of mean paired differences per metabolite and represent the effect size of change.

Overrepresentation analysis: The significant overlap between metabolite classes and stable, increasing or decreasing metabolite sets was determined using hypergeometric tests. *P*-values were corrected for multiple testing using the BH-method. The background of metabolites for hypergeometric tests was set to 680, the total number of metabolites assessed. “Partially characterised molecules” were not assessed as this category was deemed too diverse.

Correlation heatmap: The correlations between all 18 determinants were calculated with the hector function from the polychor R packages, which computes the Pearson product-moment correlations between numeric variables, polyserial correlations between numeric and ordinal variables, and polychoric correlations between ordinal variables. Categorical variables were coded to ordinal variables.

*Figure legends*

Figure 4 addition:

**B** *Significant baseline determinants for PC-baseline scores*: *(Demographic):* PC1 – sex (*p <* 0.001), age (*p <* 0.001); PC2 – sex (*p <* 0.001), age (*p <* 0.001); PC3 – sex (*p <* 0.001), age (*p <* 0.001); PC4 – age (*p =* 0.004), urbanisation (*p* *=* 0.008), education (*p <* 0.001); PC5 – age (*p* < 0.001), urbanisation (*p* *=* 0.009), education (*p =* 0.02); (*Psychosocial*): PC1 – partner status (*p <* 0.001), no high strain work (*p =* 0.006), high strain work (*p <* 0.001); PC2 – partner status (*p =* 0.006), no high strain work (*p* *=* 0.006), daily hassles (*p* *=* 0.008), childhood trauma (*p =* 0.001); PC3 – partner status (*p* < 0.001), no high strain work (*p* < 0.001), childhood trauma (*p* < 0.001); PC4 – leisure activity score (*p* < 0.001), childhood trauma (*p* *=* 0.01), recent negative life events (*p* *=* 0.03); PC5 – childhood trauma (*p <* 0.001); (*Lifestyle*): PC1 – alcohol intake (*p <* 0.001), former smoking (*p =* 0.004); PC2 – alcohol intake (*p <* 0.001), former smoking (*p =* 0.002), current smoking (*p <* 0.001); PC3 – alcohol intake (*p <* 0.001), former smoking (*p* *=* 0.009); PC4 – alcohol intake (*p <* 0.001), physical activity (*p =* 0.04); PC5 – alcohol intake (*p* < 0.001), former smoking (*p =* 0.03), current smoking (*p =* 0.03); (*Somatic health*): PC1 – N somatic diseases (*p =* 0.003), N frequent medication (*p <* 0.001), BMI (*p <* 0.001); PC2 – N somatic diseases (*p =* 0.04), N frequent medication (*p =* 0.007); PC3 – N somatic diseases (*p =* 0.004), N frequent medication (*p* < 0.001), BMI (*p =* 0.006); PC4 – N somatic diseases (*p* *=* 0.01), N frequent medication (*p <* 0.001), BMI (*p <* 0.001); PC5 – N somatic diseases (*p* *=* 0.007), N frequent medication (*p* < 0.001), BMI (*p <* 0.001); (*Mental health*): PC1-4 – n.s.; PC5 – Depression diagnosis (*p =* 0.05); *Significant baseline determinants for ΔPC scores*: *(Demographic):* ΔPC1 – sex (*p <* 0.001), age (*p <* 0.001); ΔPC2 – sex (*p <* 0.001), age (*p <* 0.001), education (*p <* 0.001); ΔPC3 – sex (p < 0.001), age (*p <* 0.001), urbanisation (*p =* 0.009); ΔPC4 – sex (*p <* 0.001), education (*p <* 0.001); ΔPC5 – age (*p* < 0.001), education (*p <* 0.001); (*Psychosocial*): ΔPC1 – High strain work (*p* *=* 0.04), daily hassles score (*p =* 0.03), recent negative life events (*p* *=* 0.04); ΔPC2 – No high strain work (*p* *=* 0.01), leisure activity score (*p =* 0.01), ΔPC3 – n.s.; ΔPC4 – leisure activity score (*p =* 0.002); ΔPC5 – No high strain work (*p <* 0.001), leisure activity score (*p =* 0.03); (*Lifestyle*): ΔPC1 – alcohol intake (*p =* 0.006), former smoking (*p =* 0.001); ΔPC2 – n.s.; ΔPC3 – alcohol intake (*p =* 0.008); ΔPC4 – alcohol intake (*p =* 0.001), current smoking (*p =* 0.03), physical activity (*p =* 0.03); ΔPC5 – alcohol intake (*p =* 0.02), current smoking (*p =* 0.005); (*Somatic health*): ΔPC1 – N frequent medication (*p =* 0.003), BMI (*p =* 0.005); ΔPC2 – N frequent medication (*p =* 0.006), BMI (*p =* 0.03); ΔPC3 – n.s.; ΔPC4 – N frequent medication (*p <* 0.001), BMI (*p <* 0.001); ΔPC5 – N somatic diseases (*p* < 0.001), N frequent medication (*p <* 0.001), BMI (*p <* 0.001); (*Mental health*): ΔPC1-5 – n.s. All *P*-values were corrected for multiple testing with the BH-method.

Figure 5 addition:

**A** *Significant baseline determinants for PC-baseline scores:* PC1 – sex (*p <* 0.001), age (*p <* 0.001), alcohol intake (*p <* 0.001), current smoking (*p =* 0.02), BMI (*p <* 0.001); PC2 – sex (*p <* 0.001), age (*p <* 0.001), daily hassles score (*p =* 0.01), current smoking (*p <* 0.001), N frequent medication (*p <* 0.001); PC3 – sex (*p <* 0.001), age (*p <* 0.001), alcohol intake (*p <* 0.001), N frequent medication (*p <* 0.001); PC4 – sex (*p =* 0.001), age (*p <* 0.001), urbanisation (*p =* 0.004), education (*p =* 0.002), leisure activity score (*p =* 0.001), childhood trauma score (*p =* 0.004), alcohol intake (*p <* 0.001), N frequent medication (*p <* 0.001), BMI (*p <* 0.001); PC5 – age (*p <* 0.001), urbanisation (*p =* 0.01), alcohol intake (*p <* 0.001), N frequent medication (*p <* 0.001), BMI (*p <* 0.001). *Significant baseline determinants for ΔPC scores:* ΔPC1 – sex (*p <* 0.001), age (*p <* 0.001), education (*p =* 0.03), daily hassles score (*p <* 0.001); ΔPC2 – sex (*p <* 0.001), age (*p <* 0.001), education (*p =* 0.04), no high strain work (*p =* 0.01), high strain work (*p =* 0.01), N frequent medication (*p <* 0.001); ΔPC3 – sex (*p <* 0.001), age (*p <* 0.001); ΔPC4 – alcohol intake (*p =* 0.001), N frequent medication (*p <* 0.001), BMI (*p <* 0.001); ΔPC5 – age (*p <* 0.001), education (*p =* 0.02), alcohol intake (*p <* 0.001), former smoking (*p =* 0.01), current smoking (*p <* 0.001), N somatic diseases (*p =* 0.005), N frequent medication (*p <* 0.001), BMI (*p <* 0.001). All *P*-values have been corrected for multiple testing with the BH-method. **B** Estimates and 95% confidence intervals from full models for PC1 to PC5, with either PC-baseline score or metabolome change PC score (ΔPC) as outcomes. Outcomes and numerical predictors have been scaled using the robust scaling method. Intercept and shipment covariate were always included but are not shown in the forest plot. For models with ΔPC as outcome the respective PC-baseline score was included as a covariate but is not shown either.
